# Spirometry parameter prediction using Acoustic characteristics of Cough

**DOI:** 10.1101/2025.04.22.25326231

**Authors:** Sujith Thomas Chandy, Balamugesh Thangakunam, Prasanna Samuel Premkumar, Devasahayam Jesudas Christopher, Gowrisree Rudraraju, Jayanthy Govindaraj, Chintakindi Lavanya Ratna Venkata, Harsha Vardhan Reddy Narreddy, Charishma Gottipulla, Shubha Deepti Palreddy, Kattula Durga Madhuri, Venkat Yechuri, Manmohan Jain, Narayana Rao Sripada

## Abstract

Spirometry evaluates lung function by measuring airflow post-maximal inspiration, using parameters like FEV1, FVC, and the FEV1/FVC ratio for respiratory disease classification and severity monitoring. Cough, by inducing intrathoracic pressure changes, reflects respiratory pathology, impacting airflow velocities and cough sound properties. A correlation exists between spirometry values representing air flow-volume properties and acoustic features of cough sounds. The present study explores the correlation between cough sounds and spirometry values (FEV1, FVC, and FEV1/FVC ratio) for assessing respiratory health. Utilizing machine learning models, trained on cough sound data labelled with corresponding spirometry pathologies, the research demonstrates, ability of swaasa in predicting spirometry values. The regression algorithm for predicting spirometry parameters, yields optimal results with FEV1/FVC prediction showing 70.47% accuracy, 77.37% sensitivity, and 68.54% specificity. FVC prediction demonstrates 66.04% accuracy, 87% sensitivity, and 48.38% specificity. The study underscores the potential of AI-based cough sound analysis for detecting respiratory abnormalities, offering a promising avenue for diagnosis in resource-limited settings.

## Introduction

Respiratory health assessment is a critical component of preventive healthcare and early diagnosis of chronic respiratory diseases is important to reduce its severity and effects. The diagnostic approach recommended by the Global Alliance against Chronic Respiratory Diseases (GARD) involves a systematic progression, with one of the crucial steps being the assessment of lung function (1). Pulmonary function tests (PFTs) enable the physicians to provide a measurable evaluation of pulmonary function in many clinical situations without the necessity for a direct physical examination of the lungs (2). In accordance with the guidelines established by the international joint task force comprising the European Respiratory Society and the American Thoracic Society (EUR/ATS), spirometry continues to be recognized as the benchmark for precise diagnosis and consistently reproducible measurement of lung function (3).

Spirometry serves as a diagnostic tool for several common respiratory conditions such as asthma and chronic obstructive pulmonary disease (COPD), while also playing a crucial role in monitoring the progression of various respiratory disorders. The key spirometry measurements include forced vital capacity (FVC), forced expiratory volume in the first second (FEV1), and the FEV1/FVC ratio. The spirometry procedure involves three phases: 1) taking a maximal breath in; 2) delivering a forceful exhalation; and 3) maintaining complete exhalation until the end of the test. Normal findings of spirometry are an FEV1/FVC ratio of greater than 0.70 and both FEV1 and FVC above 80% of the predicted value (2). Typically, a clinical significance is attributed to a 10% alteration in FEV1 from baseline over a one-year period in individuals without respiratory conditions (3).

Spirometry results are commonly juxtaposed with predicted or reference values (4). The forced expiratory ratio (FEV1/FVC) and forced vital capacity (FVC) play a crucial role in distinguishing between obstructive, restrictive, and normal breathing patterns (5). Deviations from normal values may suggest the presence of obstructive respiratory disorders like asthma and COPD, as well as restrictive conditions such as interstitial lung disease and obesity (4,6,7). Furthermore, the severity of obstructive diseases can be assessed through the measurement of FEV1.

While spirometry stands as the gold standard for diagnosing respiratory conditions, it is frequently perceived as an expensive and time-intensive process. This perception can contribute to underdiagnosis or misdiagnosis (8,9). Numerous studies highlight the underutilization or unavailability of spirometry in primary care settings and non specialized areas (10,11). In primary care, for instance, the percentage of spirometry performed according to American Thoracic Society/ European Respiratory Society (ATS/ERS) criteria is often below 50%, and there is often low agreement between general practitioners and pulmonologists on diagnoses (12). Underserved populations, including older adults and those with lower socioeconomic status, bear a disproportionate burden due to these gaps in care (13-16). Various factors contribute to these challenges; for example, spirometry involves labor-intensive and time-consuming procedures, necessitating well-trained professionals for its proper execution (17,18). Also, a majority exceeding 50% of individuals with chronic respiratory diseases reside in low and middle-income countries (19), where both medical resources and expertise are limited. This underscores the necessity for the development of new techniques and easily accessible devices capable of conducting these tests without requiring substantial medical expertise.

A significant trend expected to have a substantial impact on the future of spirometry involves the ongoing transition from in-laboratory testing to at-home testing. The current wave of affordable portable electronic spirometers is primarily designed and promoted for home use by patients. These spirometers come equipped with features like connected smartphone apps offering usage guidance and performance feedback (20-22). There are some limitations as not all devices meet the ATS/ERS quality standards nor are they suitable for all patient populations without proper education and training. Furthermore, studies that show a clear relationship between home spirometry use, clinical important outcomes and cost-effectiveness are still lacking (23,24).

Few machine learning techniques involve analyzing the “area under the expiratory flow– volume curve” to create a robust classification algorithm. This algorithm effectively distinguishes between normal respiratory patterns, obstructive diseases, restrictive diseases, and mixed impairments using spirometry alone (25).

The phases of spirometry and cough physiology share some similarities like mechanics of generating expiratory airflow, and contraction of respiratory muscles to create pressure in the airways. These similarities lead to the possibility that spirometry results can be estimated using cough sound characteristics. These innovations are proven effective in monitoring, diagnosing, and assessing treatment responses in patients with airway disorders, including asthma, chronic obstructive pulmonary disease (COPD), and even COVID-19 (26-29).

Nemati et al. (30) proposed a machine learning model that integrated standard signal processing features and domain-specific features to distinguish cough sounds obtained through spirometry. The model achieved a sensitivity of 92.38% and specificity of 90%. Kosasih et al. (31) proposed a cough detection method using a spirometry device, transmitting cough sounds to a mobile phone with (Artificial Intelligence) AI models. Employing classifiers like LR, ANN, SVM, and RF, the ANN achieved 86% sensitivity, 91% specificity, 91% accuracy, and 88% F1-score. So the results from existing literature suggests that there is correlation between spirometry parameters and cough features. And directly predicting spirometry parameter values from cough sounds provides information about restrictive, obstructive and mixed pattern related lung pathologies.

In a study done to estimate spirometry readings for asthmatic and healthy subjects using cough sounds, Rao et al. (32), The reported root mean square error (RMSE) of 0.48L, 0.57L, and 0.08 in estimating FEV1, FVC, and FEV1/FVC, respectively. Similarly, Sharan et al. utilized cough sound descriptors to build regression models, showing a strong positive correlation in predicting FEV1 and FVC and a moderate positive correlation in predicting FEV1/FVC. This suggests the feasibility of predicting spirometry results through cough sound analysis (33,34).

The present study explores the prediction of spirometry parameters using acoustic characteristics of cough. The various elements such as frequency, intensity, duration etc of the cough sounds were derived/measured using the Swaasa AI Platform (Software As a Medical Device). Cough sounds were recorded using the mobile interface of Swaasa® device. Compared to the other study (31) where the cough signals were manually segmented, the pre-processing of Swaasa device helps in automatically detecting cough sounds by eliminating all other information. As, the correlation between spirometry values reflecting air flow-volume properties of lung function was exemplified in our preceding study (34), the current study aims to identify underlying respiratory pathology due to abnormalities in the mechanical properties of cough by predicting spirometry parameters FEV1, FVC, and FEV1/FVC using cough sound analysis.

## Materials and Methods

### Data collection

This study (including the protocols and subject recruitments) was approved by the CMC (Christian Medical College) - IRB (Institutional Review Board), Vellore, Tamil Nadu, India. From every subject three audio recordings were collected before taking a spirometry test. Each record was a ten second voluntary audio cough recording. Spirometry parameter regression models were evaluated on 2506 records collected from 1202 subjects. A mobile application “Swaasa” developed by Salcit Technologies, paired with a smartphone, was used to collect the subject’s cough sounds. The smartphone was placed 5-8 cm away from the mouth of the subject at an angle of roughly around 45°. The sounds were recorded under the onsite instruction. And the sampling frequency was 44,000 Hz. Each subject was instructed to voluntarily cough at least three times within 10s with deep inspirations. Along with the cough sound, age, height, weight, and gender of the subjects were collected, and this study did not collect any personal information of the subjects.

### Event Extraction

Event extraction was carried out from the collected audio cough records using the moving window signal standard deviation technique. A cough/non-cough classifier was used to segregate the events into actual coughs and non-coughs such as silence, speech, fan sounds, vehicle sounds like horn, and noise.

### Feature Extraction

The features were extracted from the time as well as frequency domain of each cough event. The important time domain features that were taken into consideration were Zero crossing rate (ZCR) and Energy. The frequency domain features which were utilized for data analysis are MFCC, Spectral centroid, Spectral bandwidth, and Spectral roll-off 25. The features were extracted for each frame within the cough signal. Each frame was typically about 20 ms in duration. The cough event duration can vary from anywhere between 200 ms to 700 ms.

### Feature Selection

The total features extracted were 209, that includes age, gender, 120 Mel Frequency Cepstral coefficients (40 MFCC, 40 first derivatives of MFCC, 40 second order derivatives of MFCC), 9 spectral features (spectral centroid, spectral roll-off, spectral bandwidth, dominant frequency, spectral skewness, spectral kurtosis, spectral crest, spectral spread and spectral entropy), 33 chroma features (11 chroma, 11 first derivatives of chroma, 11 second derivatives of chroma), 18 contrast features (6 contrast, 6 first derivatives of contrast, 6 second derivatives of contrast), 15 tonnentz features (5 tonnentz, 5 first derivatives of tonnentz, 5 second derivatives of tonnentz), 3 Zero-crossing rate (ZCR, first derivatives of ZCR, second derivatives of ZCR), 3 Energy (Energy, first derivatives of energy, second derivatives of energy), 3 skewness (skewness, first derivatives of skewness, second derivatives of skewness), 3 kurtosis (kurtosis, first derivatives of kurtosis, second derivatives of kurtosis). On these features, we did correlation analysis and recursive feature elimination (RFE) to rank the feature according to their importance. Correlation-based feature selection was used to reduce the feature size from 209 to 170, and highly correlated features were removed to prevent overfitting and improve the performance of the model. Primary features include all the 170 features. The secondary features included age (categorized), gender, symptoms, cough type (dry/wet), and cough duration. The cough type is derived from the primary features and cough duration is derived from audio signal. Whereas, both secondary and primary features were used to train the AutoML regression model.

### Regression model flow diagram

Cough sounds can provide insights into airway dynamics and lung mechanics, which are also assessed by spirometry. The spirometry parameter readings of Forced Expiratory Volume (FEV1), Forced Vital Capacity (FVC), and the FEV1/FVC ratio are used to classify respiratory diseases and monitor disease severity. Cough expedites the generation of high intrathoracic pressure. Depending on the site of respiratory pathology, the intrathoracic pressure changes and this affects the airflow velocities and mechanical properties of the cough. There is a correlation between spirometry values reflecting air flow-volume properties of lung function and cough sound acoustic features. Table 1 highlights the differences between voluntary cough and Spirometry.

**Table 1:** Characteristics of voluntary cough and spirometry.

|  |  |  |
| --- | --- | --- |
| Feature | Voluntary cough | Spirometry |
| Purpose | Clears airways | Measures lungs function |
| Phases | Inspiration, Compression, Forced expiration | Forced Inspiration, Expiration |
| Pressure | Pressure is built for strong expulsion | Pressure is built because of breathing maneuvers |
| Usage | Cough strength/effectiveness is assessed to identify respiratory pathology. | Diagnose respiratory condition, monitor disease progression. |

The regression model for spirometry variables prediction is detailed in Figure 1. Cough sounds were recorded and sampled by a smartphone. For each single cough sound, multi-dimensional features were extracted. The time domain, frequency domain, time-frequency features were optimized using feature selection and then normalized using standard normalization. The trained result of the three pulmonary function parameters – FEV1 %, FVC% and FEV1/FVC% were used as input in the regression model.

**Figure 1:**
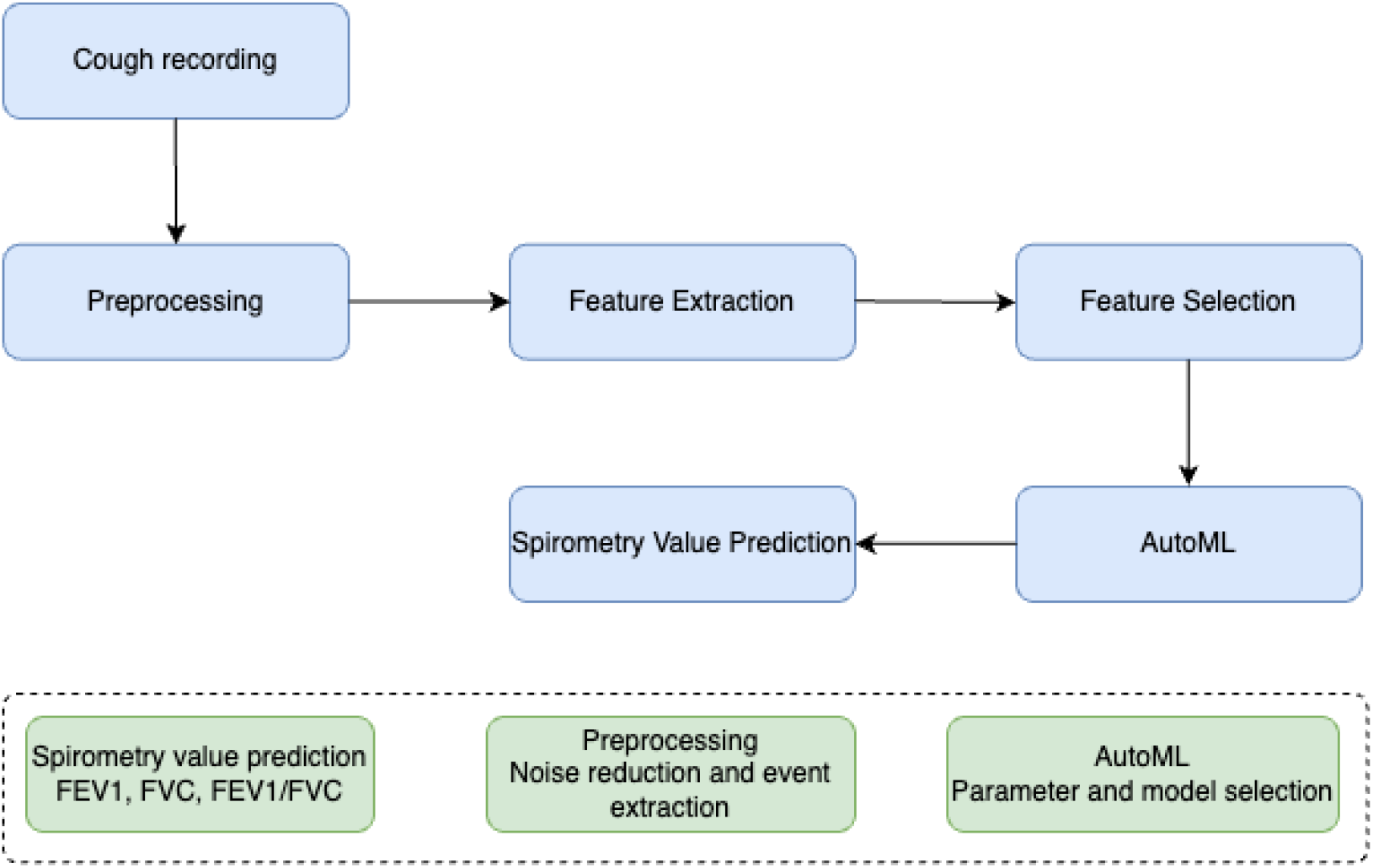
Swaasa regression model

From every subject three audio recordings were collected before taking a spirometry test. Each record was a ten second voluntary audio cough recording. AutoML (Automated machine learning), a python library from auto-sklearn, was used to build the prediction models. AutoML automatically selects the best algorithm and best parameter. After the key measurements of the spirometry parameters FEV1, FVC and FEV1/FVC are predicted using the AutoML regression model the values are classified as following

- FEV1/FVC % values are normal if the predicted value is >70% otherwise it is abnormal.
- FVC % values are normal if the predicted value is >70% otherwise it is abnormal
- FEV1 % values are normal if the predicted value is> 80% of predicted; Mild Obstruction of FEV1% values is between 65% and 79% ; Moderate Obstruction if FEV1% value is between 50% and 64% and Severe Obstruction if FEV1% value is less than 50% of predicted.

Spirometry parameter regression models were evaluated on 2506 records collected from RESULTS

### Performance of the prediction models

Table 2 illustrates the performance of regression prediction models. FEV1/FVC and FVC prediction models are binary classifiers, and FEV1 prediction model is a multiclass classifier.

**Table 2a:** Performance of FEV1/FVC% value prediction model.

|  |  | Total records | FEV1/FVC% Swaasa prediction |  |
| --- | --- | --- | --- | --- |
|  |  |  | Abnormal | Normal |
| Actual condition | Abnormal | 548 | 424 | 124 |
|  | Normal | 1958 | 616 | 1342 |

**Table 2b:**
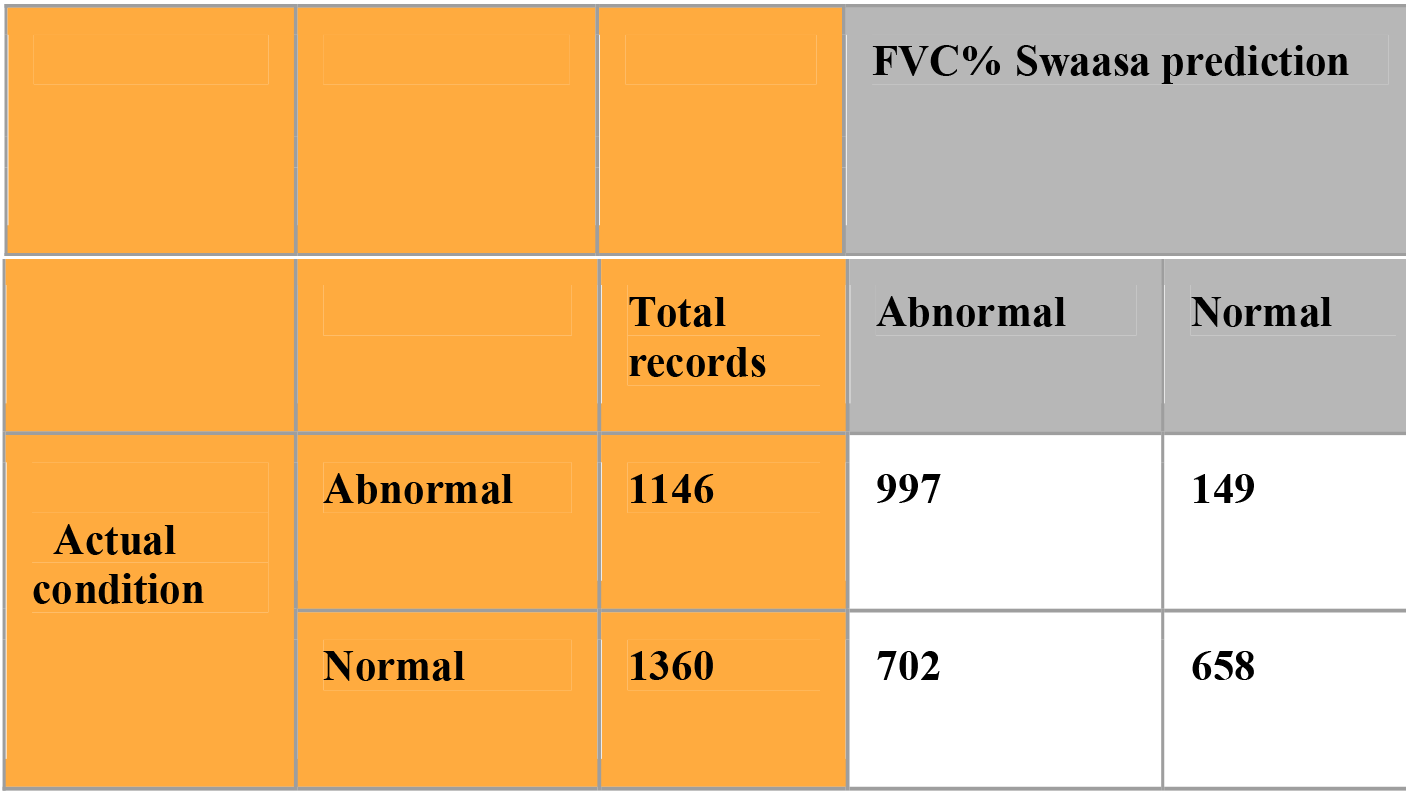
Performance of FVC% value prediction model.

**Table 2c:**
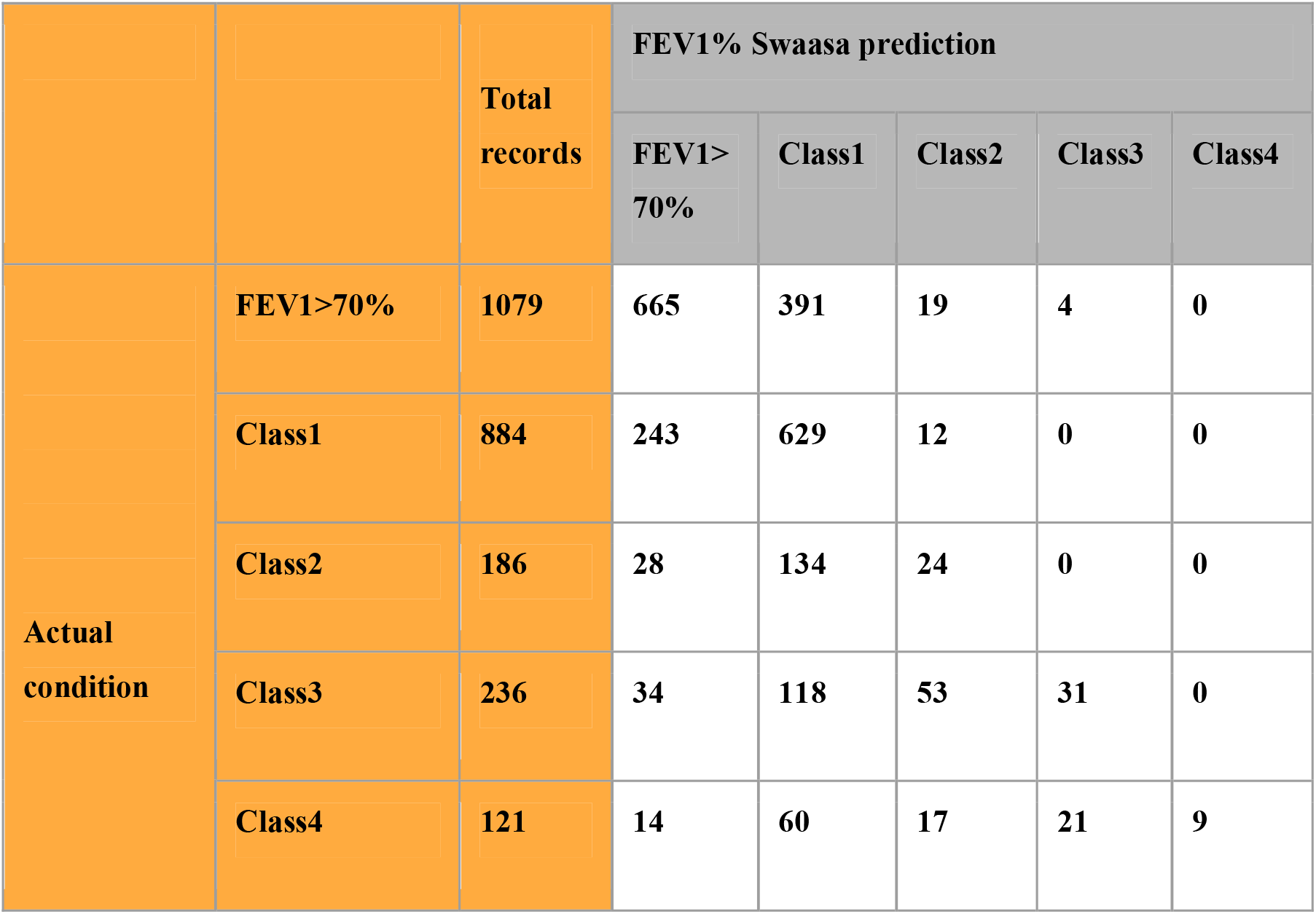
Performance of FEV1% value prediction model.

|  |  | Total records | FEV1% Swaasa prediction |  |  |  |  |
| --- | --- | --- | --- | --- | --- | --- | --- |
|  |  |  | FEV1>70% | Class1 | Class2 | Class3 | Class4 |
| Actual condition | FEV1>70% | 1079 | 665 | 391 | 19 | 4 | 0 |
|  | Class1 | 884 | 243 | 629 | 12 | 0 | 0 |
|  | Class2 | 186 | 28 | 134 | 24 | 0 | 0 |
|  | Class3 | 236 | 34 | 118 | 53 | 31 | 0 |
|  | Class4 | 121 | 14 | 60 | 17 | 21 | 9 |

### Comparison of the performance metrics of different regression models

The root mean square error (RMSE), coefficient of determination (*R*^2^), sensitivity and specificity were taken as evaluation indexes. The Auto ML regression model has selected the top three best models and we have built an embedded model with best models selected by AutoML to give most possible best results for FVE1%, FVC% and FEV1/FVC% value prediction models. Table 3 illustrates the metrics of the top three selected models for each prediction model.

**Table 3:** Performance metrics of the different regression models for each spirometry parameter

| FVC prediction model |  |  |  |  |
| --- | --- | --- | --- | --- |
|  | R2 score | RMSE | Sensitivity | Specificity |
| SVR_e = 0.01 | 0.55 | 0.52 | 0.77 | 0.42 |
| SVR | 0.56 | 0.51 | 0.79 | 0.38 |
| GBR | 0.83 | 0.31 | 0.82 | 0.72 |
| FEV1 prediction model |  |  |  |  |
|  | R2 score | RMSE | Sensitivity | Specificity |
| SVR | 0.44 | 0.53 | 0.93 | 0.25 |
| SVR_e = 0.01 | 0.43 | 0.54 | 0.94 | 0.26 |
| LR | 0.46 | 0.52 | 0.95 | 0.22 |
| FEV1/FVC prediction model |  |  |  |  |
|  | R2 score | RMSE | Sensitivity | Specificity |
| SVR_e = 0.01 | 0.23 | 15.0 | 0.77 | 0.69 |
| SVR | 0.23 | 15.0 | 0.77 | 0.69 |
| GBR | 0.28 | 14.5 | 0.78 | 0.65 |

The three models selected for each prediction model are as follows -

- FEV1% prediction models are Support Vector Regression (SVR); SVR = 0.01 regression (where a specific parameter, often called C, is set to 0.01); Gradient Boosted Regression (GBR).
- FVC% prediction models are Support Vector Regression (SVR); SVR = 0.01 regression (where a specific parameter, often called C, is set to 0.01); Linear Regression (LR)
- FEV1/FVC% prediction models are Support Vector Regression (SVR); SVR = 0.01 regression (where a specific parameter, often called C, is set to 0.01); Gradient Boosted Regression (GBR)

### Effect of biological attributes

Regression models were analysed at different level –

1. Overall classification
2. Classification based on age groups
3. Classification based on gender
4. Classification based on BMI

#### Spirometry Value Prediction

Based on a correlation established between the spirometry values (FEV1, FVC, and FEV1/FVC), Swaasa predicted its values using cough as the input Swaasa outputs numerical values for FEV1%, FVC%, and (FEV1/FVC) %.

#### FEV1 prediction Class

This refers to the classification of the FEV1 % predicted values derived from spirometry testing. The categorizations align with FEV1 % predicted ranges as outlined: Mild (>70), Moderate (69 – 50), Severe (35 - 49), Very Severe (<35).

#### FVC prediction Class

**T**his refers to the classification of the FVC % predicted values derived from spirometry testing. The categorizations align with FVC % predicted ranges as outlined: Normal (>70), Abnormal (<70).

#### FEV1/FVC prediction Class

**T**his refers to the classification of the FEV1/FVC % predicted values derived from spirometry testing. The categorizations align with FEV1/FVC % predicted ranges as outlined: Normal (>70), Abnormal (<70).

Table 4 and table 5 details the binary and multi classification for spirometry parameters

**Table 4a.**
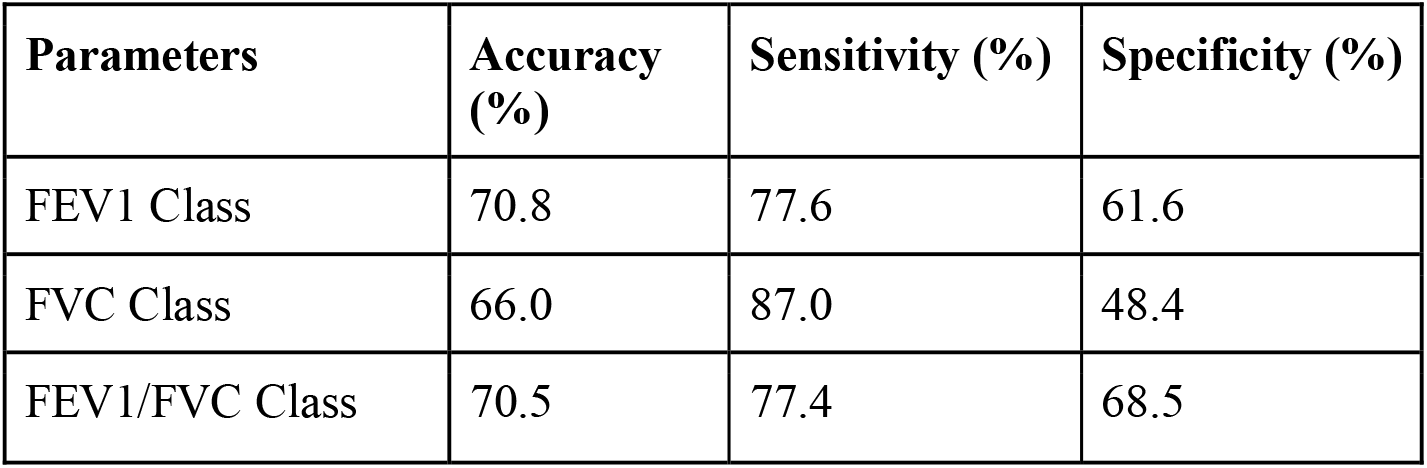
Confusion Matrix Statistics for spirometry variables with binary classification

| Parameters | Accuracy (%) | Sensitivity (%) | Specificity (%) |
| --- | --- | --- | --- |
| FEV1 Class | 70.8 | 77.6 | 61.6 |
| FVC Class | 66.0 | 87.0 | 48.4 |
| FEV1/FVC Class | 70.5 | 77.4 | 68.5 |

**Table 4b.**
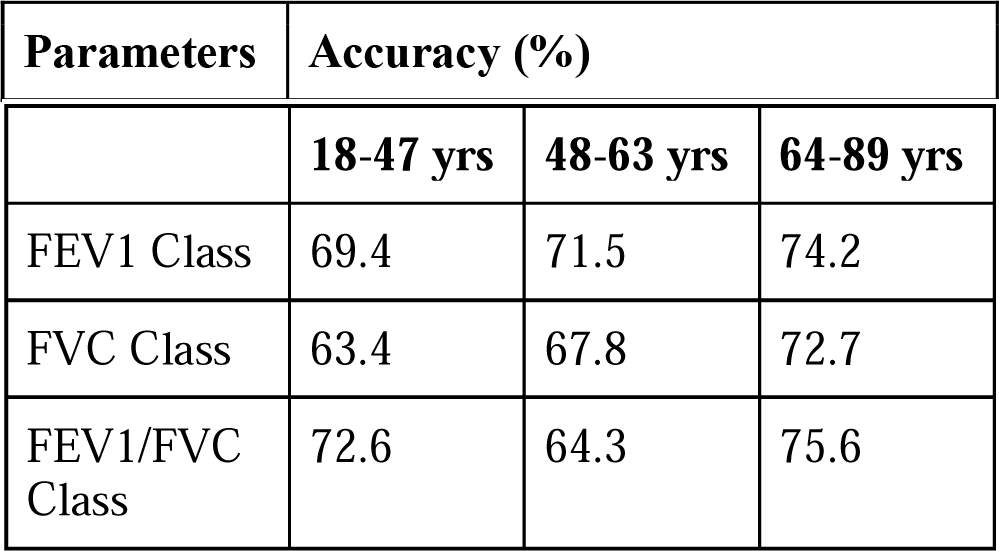
Confusion Matrix Statistics for spirometry variables with binary classification based on Age subgroups

|  | **18-47 yrs** | **48-63 yrs** | **64-89 yrs** |
| --- | --- | --- | --- |
| FEV1 Class | 69.4 | 71.5 | 74.2 |
| FVC Class | 63.4 | 67.8 | 72.7 |
| FEV1/FVC Class | 72.6 | 64.3 | 75.6 |

**Table 4c.** Confusion Matrix Statistics for spirometry variables with binary classification based on Gender subgroups

| **Parameters** | **Accuracy (%)** |  | **Kappa Statistic (%)** |  |
| --- | --- | --- | --- | --- |
|  | **Male** | **Female** | **Male** | **Female** |
| FEV1 Class | 71.2 | 70.0 | 40.2 | 38.8 |
| FVC Class | 66.4 | 65.5 | 34.4 | 33.5 |
| FEV1/FVC Class | 66.0 | 77.9 | 33.0 | 29.7 |

**Table 4d.** Confusion Matrix Statistics for spirometry variables with binary classification based on BMI subgroups

| **Parameters** | **Accuracy (%)** |  |  |  |
| --- | --- | --- | --- | --- |
|  | **<18.5** | **18.5 – 22.9** | **23 – 24.9** | **>=25** |
| FEV1 Class | 70.7 | 72.7 | 62.3 | 74.1 |
| FVC Class | 58.7 | 64.7 | 60.8 | 70.1 |
| FEV1/FVC Class | 71.4 | 70.6 | 65.3 | 65.1 |

**Table 5.** Confusion Matrix Statistics for spirometry multi-class variables

| **Parameters** | **Accuracy (%)** | **Sensitivity (%)** | **Specificity (%)** |
| --- | --- | --- | --- |
| **FEV1 Class** |  |  |  |
| Normal | 70.6 | 61.6 | 77.6 |
| Mild | 61.8 | 71.2 | 56.7 |
| Moderate | 89.5 | 12.9 | 95.6 |
| Severe | 90.8 | 13.1 | 98.9 |
| Very Severe | 95.5 | 7.4 | 100 |

## Conclusion

Cough sound carries information related to lung health conditions. In this study, the regression prediction models (i.e., SVR, GBR, LR) are combined to predict pulmonary function parameters from voluntary cough sounds. The experimental results show that biological attributes like age, sex and weight have a minimal estimating effect on prediction models.

Pulmonary function testing is the gold standard for evaluating lung function, but it cannot meet the daily monitoring needs of patients with lung diseases. It is normal for problems to occur during the examination process. The process of coughing has something similar to pulmonary function testing, and the cough sound contains rich respiratory information, which can be determined easily. For cough sounds and pulmonary function parameters, we evaluate cough sounds by analyzing the relationship between cough sound and pulmonary function parameters. It is expected to gain its new clinical and disease control value in the diagnosis of respiratory diseases, so as to facilitate the daily lung function examination of patients with chronic respiratory diseases, and reduce the delay in timely diagnosis.This technology has great potential, and in the future, it can provide practical basis and reliable technology for large-scale screening of possible unknown infectious diseases. However, this paper only provides a preliminary discussion, there must be shortcomings, and need to be further improved. It is worth emphasizing that this study is only used as an auxiliary method for medical diagnosis and evaluation. Due to the complexity of the disease, the diagnosis process can not be limited to this technique.

## Data availability statement

Due to the nature of this research, participants of this study did not agree for their data to be shared publicly. However, the detailed analysis can be shared by the author “NRS” upon reasonable request.

## Ethical clearance

The study was registered under Clinical Trials Registry-India (CTRI/2021/04/032742) and was begun after getting the approval [Ethics Approval number - IRB Min. No. 13566 (DIAGNO)] from the CMC-IRB (Institutional Review Board).

## Author Contributions

STC and BT defined study protocols, including study design and methodology. NRS conceptualized the idea of using cough sounds for screening and diagnosing COVID-19. GR performed literature review and data analysis. BM, HVR, SDP and NKRB were involved in device development. VY and MJ created value propositions for the device. SS assisted in executing the project at Christian Medical College by providing all the resources and extending research capabilities. CG and GR performed data analysis, sample size estimation and result analysis. KLPK, SS, NJ, VSP, ST and SV provided subject matter expertise. GR and JG wrote the manuscript. All the authors provided intellectual inputs and helped in preparing the manuscript.

## Acknowledgements

This study is supported by Biotechnology Industry Research Assistance Council (BIRAC) [grant number - BT/BIPP1335/BIPP-49/20]. This is a commissioned research report on commercial terms between C-CAMP-FCDO and the UK Government (British High Commission, New Delhi). We would also like to acknowledge the team from Christian Medical College, Vellore, India for all the support provided.

## Patient and Public Involvement

Patients or the public WERE NOT involved in the design, or conduct, or reporting, or dissemination plans of our research.

## Competing interest statement

The authors have no conflicts of interest to declare. All co-authors have seen and agree with the contents of the manuscript and there is no financial interest in reporting. We certify that the submission is original work and is not under review at any other publication.

## Notes

### Competing Interest Statement

The authors have declared no competing interest.

### Clinical Trial

CTRI/2021/04/032742

### Author Declarations

The study was registered under Clinical Trials Registry- India (CTRI/2021/04/032742) and was begun after getting the approval [Ethics Approval number - IRB Min. No. 13566 (DIAGNO)] from the CMC- IRB (Institutional Review Board).

### Summary of Updates

Added table 1 and figure 1.

